# The Potential Sensor: Design and development of a novel patient-friendly instrument for non-invasive erectile dysfunction diagnostics

**DOI:** 10.1101/2025.03.27.25322960

**Authors:** Hille J. Torenvlied, Jorien T.W. Berendsen, Julian G.J. Klep, Loes I. Segerink, Jack J.H. Beck

## Abstract

This study introduces The Potential Sensor, a proof-of-principle system designed for nocturnal erection detection. The article introduces a novel approach to non-invasive ED diagnostics, which aims to optimize patient comfort and consequently system validity through avoidance of applying pressure-based measurements. A literature review on erection physiology and applicable sensor technologies showed evaluated concepts for measuring penile erection, including temperature, circumference, movement, blood oxygenation, and arterial pulse as suitable for application in a patient-friendly diagnostic system. This guided the development of system requirements for a device designed to quantify both erection duration and erection quality through simultaneous assessment of multiple physiological principles. The proposed system integrates four commercially available sensor types (thermistors, stretch sensors, accelerometers, and pulse oximeters) and connects these to a microprocessor that transmits data via Bluetooth. Initial testing of the sensor components confirmed precision and accuracy of the device, demonstrating readiness for future feasibility and clinical validation studies. The Potential Sensor offers a unique diagnostic approach, which presents as a promising alternative to traditional nocturnal penile tumescence and rigidity tests, by enhancing patient comfort. This novel approach has the potential not only to reintroduce non-invasive diagnostics in clinical practice, but also to improve the understanding of physiological mechanisms underlying erectile dysfunction.

## Introduction

Non-invasive advanced diagnostics for erectile dysfunction (ED) primarily aim to differentiate psychogenic from organic causes by assessing nocturnal erections [1]. Since its introduction in 1985, the RigiScan^®^ has been the gold standard for nocturnal penile tumescence and rigidity (NPTR) testing in ambulatory setting [2]. However, its outdated hardware and software have diminished its clinical utility [3]. In contrast, hospital-based alternatives, such as penile duplex doppler ultrasound (PDDU) following intracavernosal injection of prostaglandin E1, are invasive, underscoring the pressing need for a modern, patient-friendly alternative for non-invasive ED diagnostics [4].

Normal erectile functioning during NPTR testing suggests psychogenic ED. NPTR testing focuses on two critical indicators: erection duration and erection quality. A normal nocturnal erection defined by a penile circumference increase of 2 – 3 cm sustained for at least 10 minutes [5]. Erection quality is deemed sufficient for penetration if rigidity levels exceed 60% [5]. The RigiScan^®^ assesses rigidity using a spring constant mechanism, applying pressure (2.8N) to the penile tissue at a frequency of 20 times per hour during flaccid phases and twice per minute during erectile phases to measure tissue deformation [2]. However, this reliance on external stress can disrupt sleep quality and result in absence of rapid-eye-movement (REM) sleep periods, which raises concerns about its diagnostic validity [6].

Recent advancements in wearable devices for at-home diagnostics offer a promising pathway to modernize nocturnal erection detection. Ideally, such a system should assess erection duration and erection quality without relying on pressure-based components. Edgar et al. [7] proposed physiological principle such as penile temperature, saturation, and arterial pulse measurements. Previous proof-of-principle studies suggest that penile temperature monitoring can detect erection onset, but assessing both erection duration and quality requires integrating multiple physiological principles into the system [8]. This study introduces a novel proof-of-principle sensor system for non-pressure-based nocturnal erection detection and evaluates its readiness for future validation studies.

## Materials and Methods

This study was conducted at the University of Twente, Enschede, the Netherlands in collaboration with the Department of Urology at St. Antonius Hospital, Nieuwegein, the Netherlands. A comprehensive literature review was performed to identify physiological principles for a patient-friendly diagnostic sensor system (“I. System Conceptualization”). Sources included Google Scholar, PubMed, and Scopus. Based on identified concepts, system requirements were defined to propose a multi-principle design capable of non-invasively assessing erection duration and erection quality (“II. System Design”). The sensor system was subsequently developed by selecting commercially available components (III. System development”). Each component underwent a validation process to ensure accuracy and precision, enabling future feasibility and validation studies (“IV. System assessment”). For the validation study that involved human subjects, ethical approval (W25.043) was received from the Medical research Ethics Committees United, Nieuwegein, the Netherlands.

## Results

### I. SYSTEM CONCEPTUALIZATION

The Potential Sensor aims to evaluate both erection duration and quality [4,5]. Determining erection duration requires identification of both tumescence and detumescence. Penile temperature measurements have demonstrated feasibility for detecting tumescence [9-11]. Penile temperature is regulated by heat conduction from the arteria cavernosa and the corpora cavernosa towards the penile skin [12]. During erection, vasocongestion – swelling of the corpora cavernosa due to increased arterial blood supply and reduced venous outflow – reduces the distance for heat transfer, resulting in an increased penile heat flux and a rise in penile skin temperature [9]. However, penile temperature alone is insufficient for assessing detumescence, as peak temperature persists under blankets post-erection [8]. Seeley et al. [9] demonstrated that abdominal temperature decreases during tumescence. Therefore, an increase in abdominal temperature would indicate detumescence and thus allow for measurement of erection duration. Further research is required to validate this hypothesis.

To overcome the limitation of penile temperature measurements, additional physiological principles were integrated into the novel approach. Vasocongestion also induces changes in penile circumference and movement relative to the body, which are minimally influenced by environmental conditions. These movements can be effectively detected using accelerometry. Circumference changes, traditionaly measured by the RigiScan’s ring-shaped pressure sensor, could potentially be assessed in a non-pressure manner using a semi-circular, non-restraining stretch sensor.

Monitoring penile tissue oxygenation may also offer a valuable method for assessing erection duration [7]. During erection, the trapping of blood in the corpora cavernosa leads to a decline in blood oxygen levels (SpO_2_), as the surrounding tissue continues to consume oxygen [9]. In cases of low-flow priapism, penile pO_2_ levels can drop below 30 mmHg, compared to normal values above 90 mmHg [5,13]. Given that a typical nocturnal erection lasts approximately 25 minutes, SpO_2_ levels in the corpora cavernosa are expected to decrease during these episodes. Studies have confirmed the presence of SpO_2_ changes during (nocturnal) erections through pulse oximetry [14]. Consequently, monitoring penile oxygen saturation could aid in detecting both tumescence and detumescence, enabling precise determination of erection duration. Additionally, if a correlation between SpO_2_ levels and erection rigidity can be established – a relationship not yet explored– this approach could facilitate the assessment of both erection duration and erection quality.

Erection quality is traditionally assessed through penile rigidity measurements, which rely on a pressure component. As a non-pressure-based alternative, arterial pulse measurements present a theoretically robust method. Blood supply to the corpora cavernosa via the arteria cavernosa creates an increased volume difference between systole and diastole, amplifying the penile pulse. This phenomenon has been documented in measurements of the penile pulse from the arteria dorsalis [15]. During nocturnal erection, the filling of the corpora cavernosa elevates intracavernosal pressure to systolic levels, resulting in full penile rigidity [16]. When external pressure equals systolic pressure, vasoconstriction occurs, leading to the absence of an arterial pulse. This principle, analogous to the mechanism of blood pressure measurement with a sphygmomanometer, is expected to manifest in the arteria cavernosa during a fully rigid erection. Thus, detecting the absence of an arterial pulse could provide a reliable method for quantifying erection quality.

In conclusion, the assessment of erection duration and quality appears theoretically achievable through the development of a sensor system that integrates measurements of (a) penile temperature, (b) circumference, (c) acceleration, (d) blood oxygenation, and (e) arterial pulse.

### II. SYSTEM DESIGN

System requirements were defined for each of the five sensor concepts identified as potential methods for assessing erection duration and quality. Table 1 provides an overview of these requirements.

**Table 1:** System requirements of sensor components integrated into The Potential Sensor.

| Sensor Concept | System Requirements |
| --- | --- |
| *Temperature* | - Lightweight and reusable design<br>- Thermistor type sensor<br>- Small size for distal penile placement<br>- Disk shape with external temperature insulation<br>- Penile, thigh, and abdominal sensors<br>- Measurement range: 20°C to 45°C |
| *Circumference* | - Lightweight and reusable design<br>- Stretch sensor<br>- No resistance components<br>- Measurement range: 45mm to 80mm |
| *Accelerometry* | - Lightweight and reusable design<br>- Gravitational accelerometer<br>- Penile and abdominal sensors<br>- Measurement range: -2G to 2G with $G=9.81\text{m/s}^2$ |
| *Blood oxygenation* | - Lightweight and reusable design<br>- Pulse oximeter sensor<br>- Use of red and IR lights<br>- Measurement range: 60 to 100%<br>- Penetration depth of red and IR lights $\geq 5\text{mm}$ |
| *Arterial pulse* | - Lightweight and reusable design<br>- Pulse oximeter sensor<br>- Use of IR light<br>- Penetration depth of IR lights: $\geq 14\text{mm}$ |

#### Temperature measurement

Previous studies on penile temperature measurements utilized either thermistor sensors or infrared (IR) thermometry, with IR thermometry offering superior patient comfort due to its non-contact nature. However, because penile temperature changes during erection are not uniform, maintaining consistent sensor placement is critical for accurate measurements [10]. Thermistor sensors, favored for their compact size, precision, and sensitivity, can provide reliable measurements when securely attached to the skin using adhesive plasters. The most significant temperature increase during erection occurs at the glans [11]. Nevertheless, positioning the temperature sensor on the neck of the penis, just proximal to the glans, is recommended. This location accommodates foreskin mobility and ensures inclusivity for circumcised individuals [8].

In addition to the penile temperature sensor, two auxiliary thermistors should be incorporated into The Potential Sensor. A prior study suggested placing a reference temperature sensor on the thigh to distinguish between tumescence and frictional energy generated by body movement [8]. An abdominal temperature sensor is also proposed to assess detumescence and serve as a reference for body movement [9]. If abdominal temperature reliably differentiates these parameters, the thigh thermistor could be omitted from the device design, simplifying its configuration.

#### Circumference and accelerometry measurement

A study of 4,685 young Italian men reported an average increase in penile circumference from 9.59 cm to 12.03 cm during erection [17]. By placing a semi-circular stretch sensor slightly distal to the temperature sensor, these circumference changes can be measured while maintaining patient comfort. Distal placement also captures the highest acceleration, enabling the accelerometer to be co-located with the stretch sensor. The accelerometer, measuring three-dimensional accelerations, facilitates the detection of penile movements. Nocturnal erections occur during the REM sleep, which is marked by muscle atonia [18]. To distinguish between body and penile movements, a reference accelerometer should be included alongside the abdominal temperature sensor. This configuration enhances the system’s ability to accurately attribute movements to penile activity.

#### Blood oxygenation and arterial pulse measurement

Pulse oximetry sensors are commonly utilized in clinical practice for their accuracy and cost-effectiveness in measuring SpO_2_ and arterial pulse. These sensors operate using red and near-IR light, typically at wavelengths of 660 nm and 880 nm, achieving penetration depths ranging from 1 to 8 cm [19]. Clinical studies have demonstrated a penetration depth of approximately 5 mm for red light in skin and subcutaneous tissue [20]. In-house experiments conducted during PDDU examinations at St. Antonius Hospital (NL) revealed an average skin-to-corpora cavernosa distance of 5.3 mm in the flaccid state and 3.7 mm in the erect state {unpublished data}. These findings indicate that measuring SpO_2_ in the corpora cavernosa during erections is theoretically feasible. For arterial pulse measurement, the distance from the penile skin to the arteria cavernosa was found to require penetration depths of 7.8 and 14 mm for flaccid and erect states, respectively. Based on the study by Kovacsova et al. [19], which reported penetration depths between 1 and 8 cm, IR light appears capable of measuring the arterial pulse of the arteria cavernosa. To ensure stable placement near the corpora cavernosa, the pulse oximeter should be positioned alongside the penile temperature sensor on the dorsal side of the penis. This placement optimizes sensor stability and proximity to the target tissue for accurate measurements.

### III. SYSTEM DEVELOPMENT

Based on the established system requirements, various commercially available sensors were evaluated, and a suitable sensor was selected for each concept:

- **Temperature Sensor**: Circular and insulated 203AT-2 Semitec thermistors (Semitec, Tokyo, Japan) [21]
- **Stretch Sensor**: Conductive rubber cord stretch sensor (Adafruit, New York City, USA) [22]
- **Accelerometer**: ADXL345 3-axis accelerometer (TinyTronics, Eindhoven, NL) [23]
- **Pulse Oximeter**: MAX 30102 reflective pulse oximeter sensor (TinyTronics, Eindhoven, NL) [24]

These sensors were integrated into The Potential Sensor. The five sensor concepts are distributed across three components, as shown in Figure 1. All sensors are connected to an Arduino Nano 33 BLE Sense Rev2 microprocessor (TinyTronics, Eindhoven, NL), housed within the abdominal component of the system [25]. Data is wirelessly transmitted via Bluetooth Low Energy to an external storage device, such as a smartphone, placed on the patient’s nightstand. To ensure data storage for at least ten hours, a battery with a capacity of at least 600mAh was required. An 800mAh 3.7V LiPo battery (Droneshop.nl, Den Bosch, NL), measuring 45×25×9 mm and weighing 20 grams, was selected [26]. This lightweight, rechargeable battery satisfies the system’s power requirements while maintaining portability.

**Figure 1:**
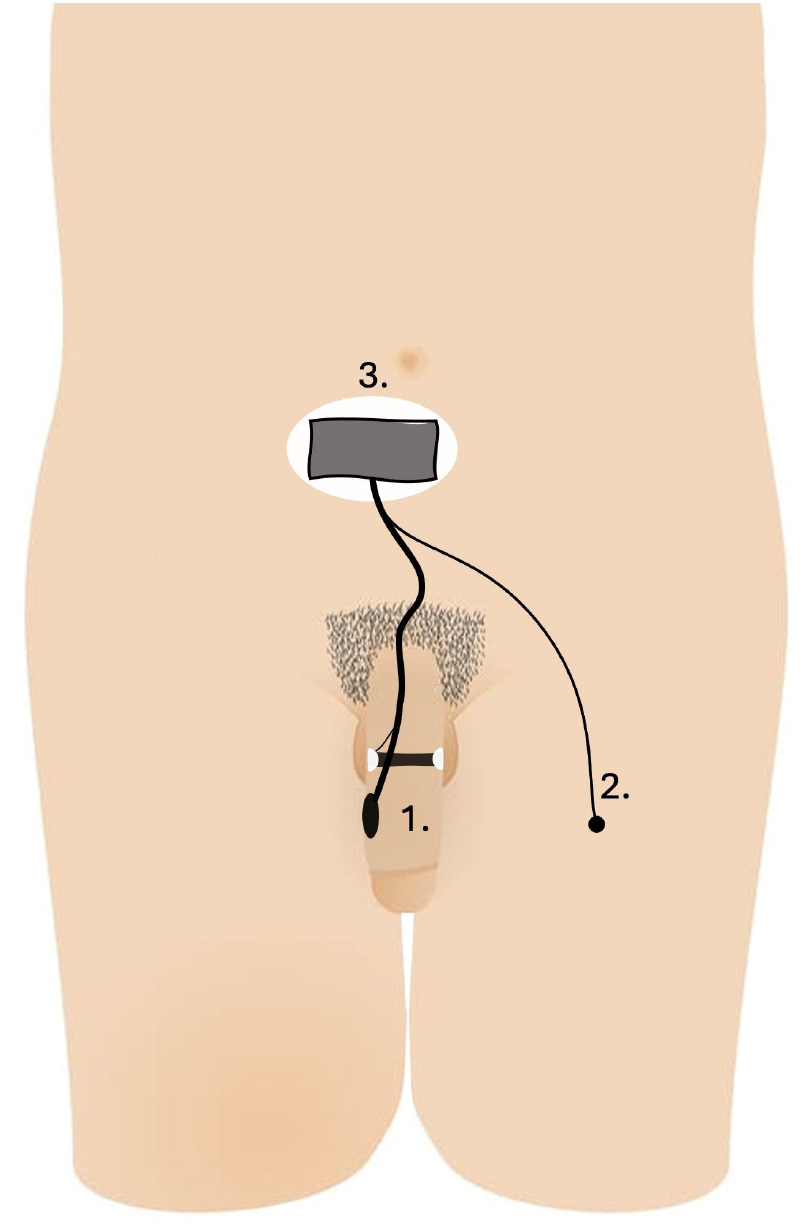
Schematic representation of The Potential Sensor system, illustrating the overall system design including: (1) the penile component containing thermistor, stretch sensor, accelerometer, and pulse oximeter, (2) the thigh thermistor, and (3) the abdominal component containing thermistor, accelerometer, microprocessor, and battery.

### IV. SYSTEM ASSESSMENT

The accuracy and precision of each sensor component were assessed, with the validation setup illustrated in Figure 2.

**Figure 2:**
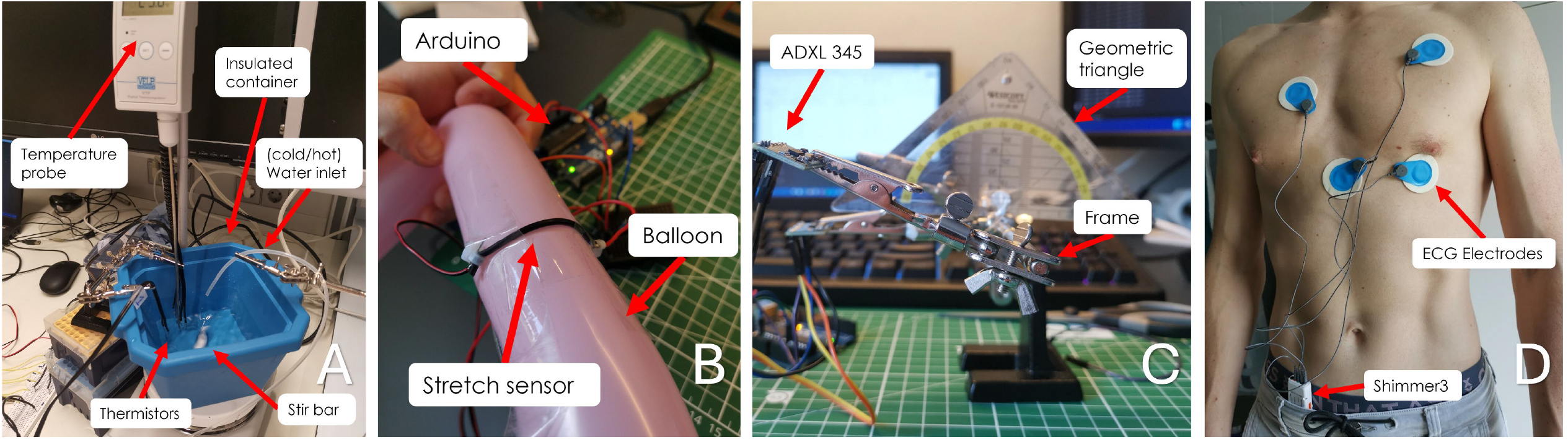
Experimental setups for system assessments: (A) Temperature Sensor, (B) Stretch Sensor, (C) Accelerometry, and (D) Pulse Oximeter.

#### Temperature sensor

The three thermistor sensors were tested in an insulated water container placed on a heating element, which included a stir bar to maintain uniform temperature distribution. A high-precision thermometer served as the reference sensor. The results, shown in Figure 3, demonstrated a linear resistance-to-temperature correlation of -282.81Ω/°C with a coefficient of determination (R^2^) of 0.937, indicating sufficient accuracy and precision for clinical use.

**Figure 3:**
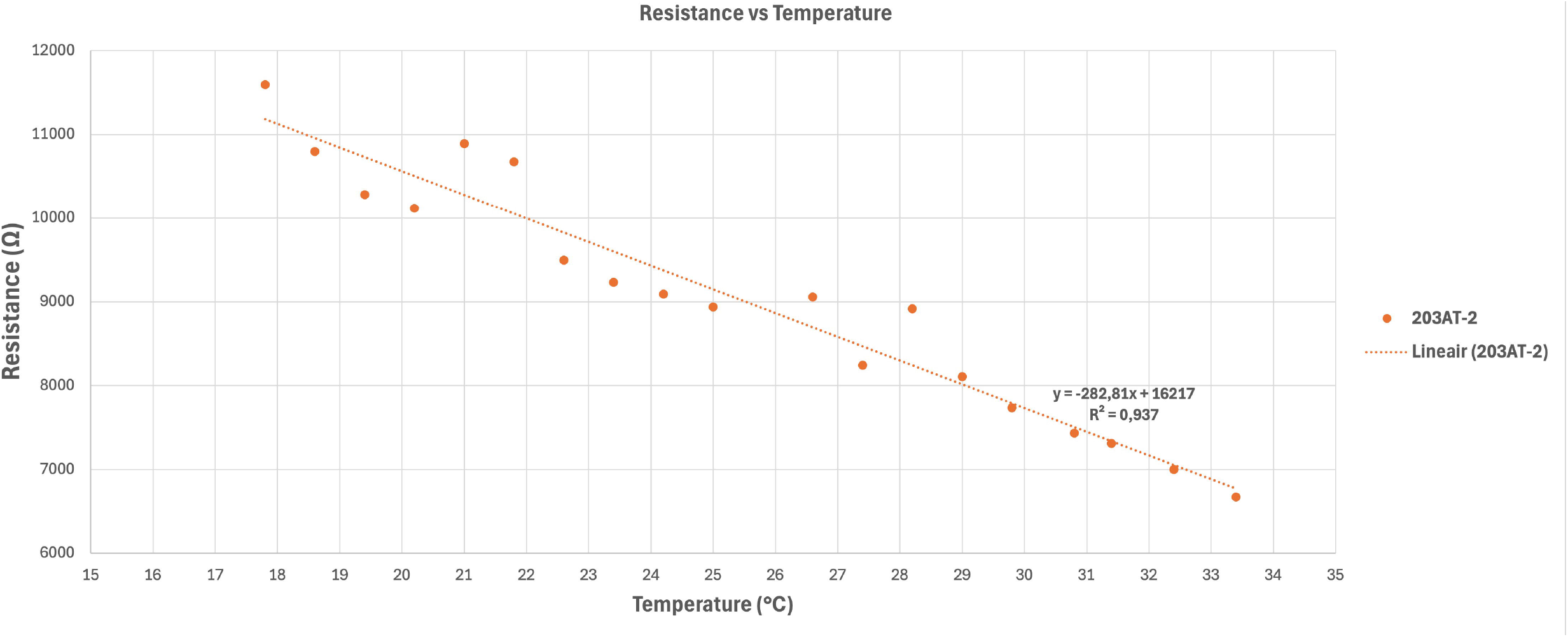
Results of the temperature sensor assessment, demonstrating a linear correlation between measured resistance and water temperature. The linear relationship is characterized by a resistance-to-temperature correlation of -238.69Ω/°C with a precision of 0.990.

#### Stretch sensor

Initial validation of the stretch sensor was performed on a stable surface by incrementally stretching the sensor by 0.5cm between 4.5 and 8.5cm. The results, presented in Figure 4, revealed a linear resistance-to-stretch correlation of -52.00Ω/cm with a R^2^ of 0.969. Further validation involved measuring variable circumferences around a modeling balloon, confirming the sensor’s capacity to detect circular changes, simulating its application within The Potential Sensor.

**Figure 4:**
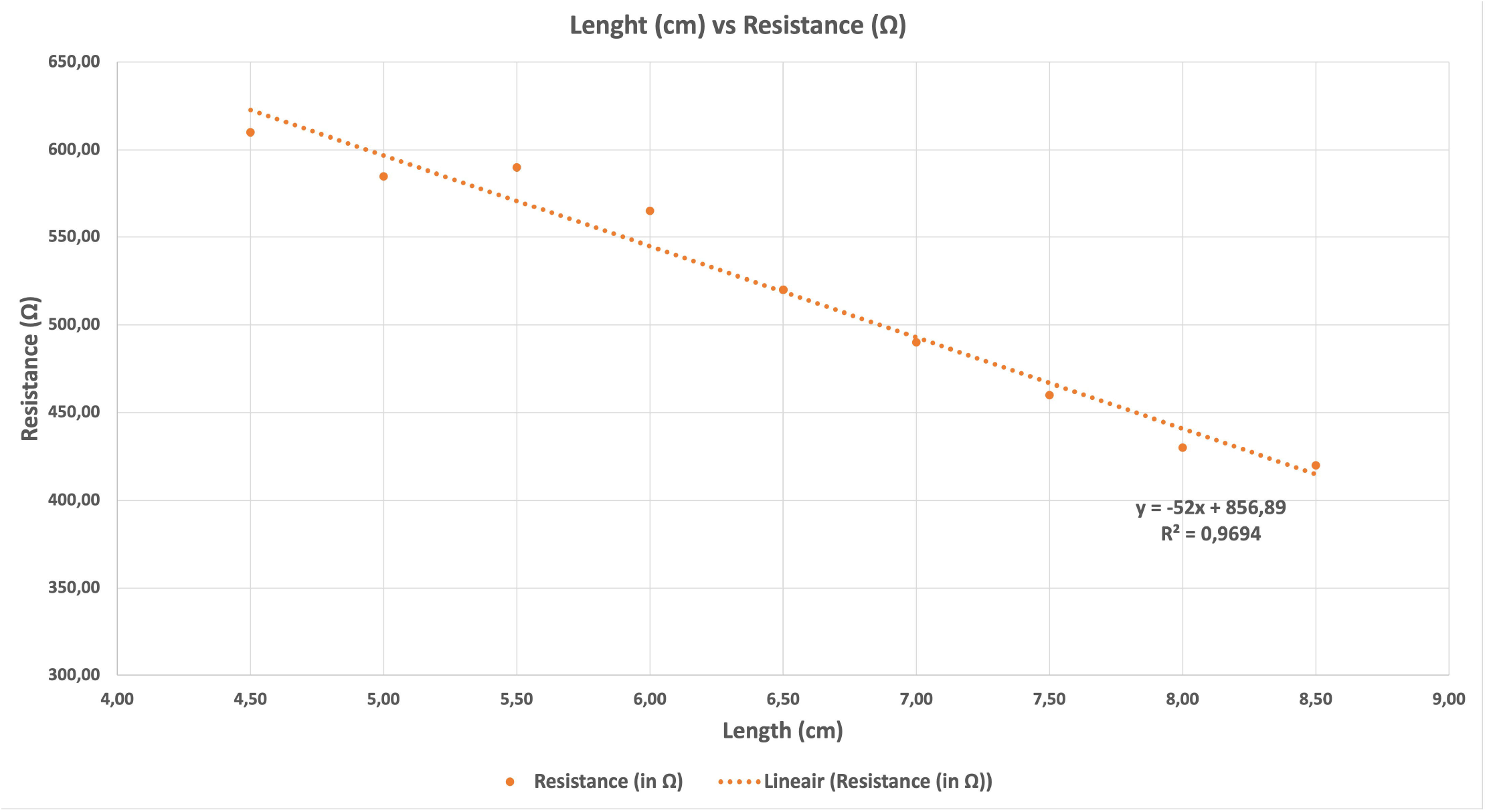
Results of the stretch sensor assessment, showing a linear correlation between changes in resistance and stretch length with a precision of 0.969.

#### Accelerometer

Validation of the penile and abdominal accelerometers was conducted using a setup that incrementally changed the sensor’s angle by 10 degrees along the Y-roll and X-roll axes to assess the detection accuracy in each direction. As shown in Figure 5, the sensor effectively detected acceleration changes, confirming its capability to monitor penile movement.

**Figure 5:**
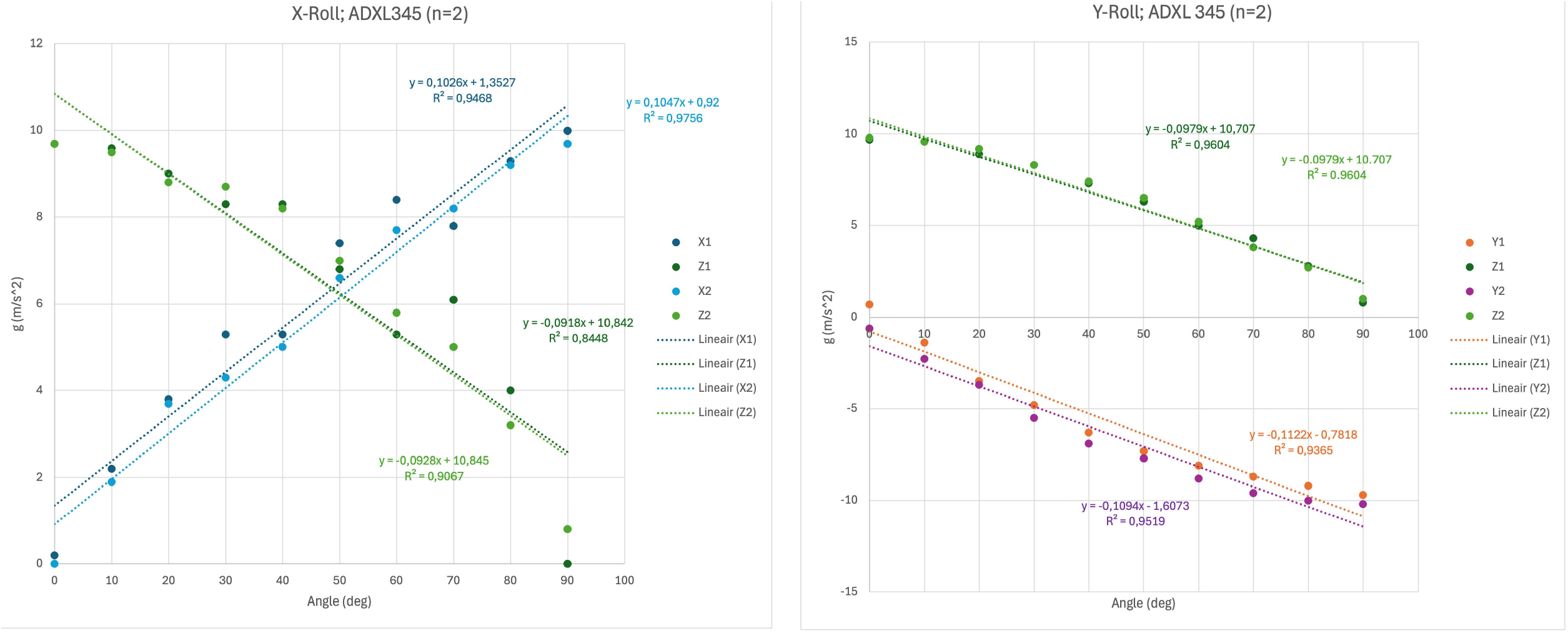
Results of the accelerometer assessment for the two ADXL345 3-axis accelerometers. The X-roll represents rotation around the X-axis (side-to-side tilting), while the Y-roll indicates rotation around the Y-axis (forward and backward tilting). X1, Y1, and Z1 correspond with the three measurement directions of sensor 1. The data show consistent linear relationships between angular rotation and accelerations, with precision values ranging from 0.844 – 0.976.

#### Pulse oximeter

The pulse oximeter’s validation for measuring arterial pulse was performed by assessing the arteria digitalis palmaris. Simultaneously, heart rate data were collected using a 3-lead Shimmer 3 Consensys ECG (Shimmer, Dublin, Ireland) [27]. The results demonstrated consistent pulse detection between the two systems, as illustrated in Figure 6.

**Figure 6:**
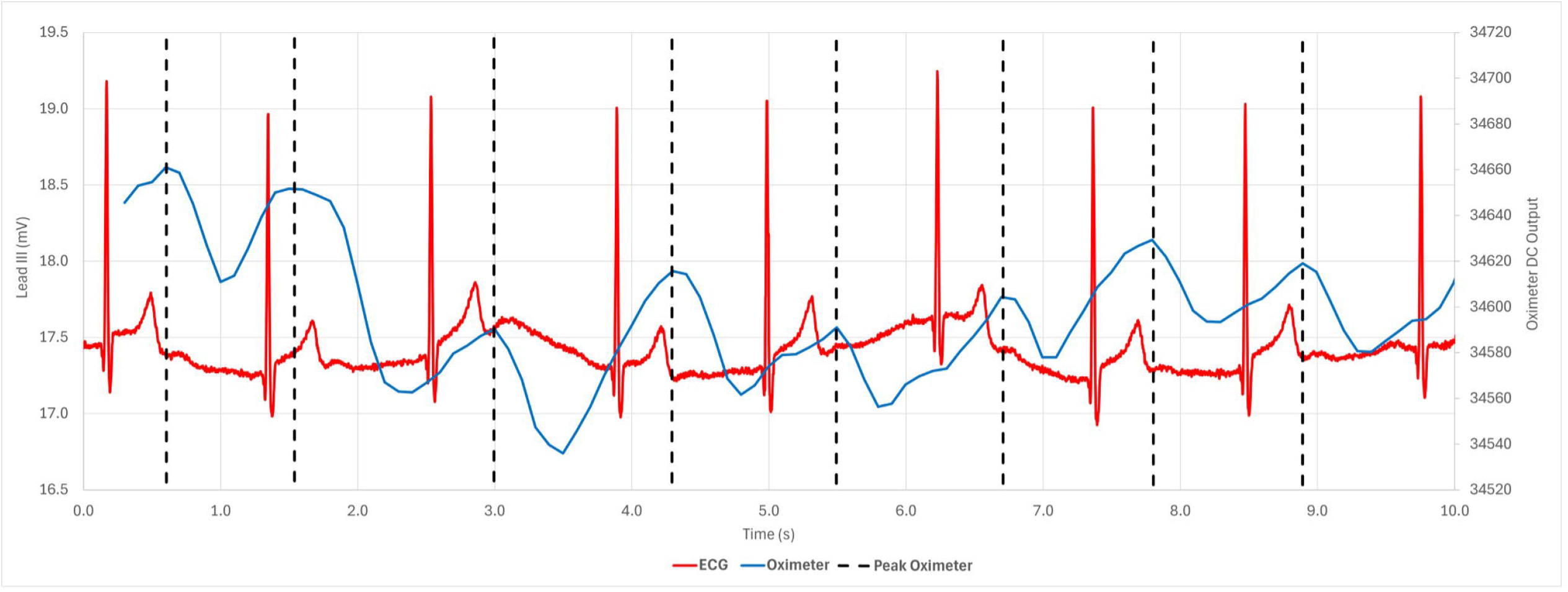
Results of the arterial pulse validation, illustrating the correlation between QRS complex of the ECG signal (red) and the peaks (black) detected by the MAX 30102 (blue). A physiological delay of approximately 0.25 seconds is observed at a heartrate of 55 beats/minute for both measurements. Variations in peak height of the MAX 30102 are due to movement artefacts.

## Discussion

‘The Potential Sensor’ is a proof-of-principle system designed to present a novel approach to nocturnal erection detection. Unlike current NPTR tests that rely on pressure-based component, The Potential Sensor offers a patient-friendly alternative. This system represents the first modernized, non-invasive diagnostic tool for advanced ED diagnostics that does not use pressure-based measurements, which are often linked to patient discomfort. By integrating measurements of penile temperature, circumference, accelerometry, blood oxygenation, and arterial pulse, The Potential Sensor theoretically enables the assessment of both erection duration and quality. The four selected sensors have demonstrated sufficient precision, supporting their potential use in future feasibility studies and clinical validation.

In recent years, several researchers and companies have developed modernized systems for nocturnal erection detection, such as the ‘EDSEN sensor’ (Sng et al. [28]), the ‘ADAM sensor’ (Konstantinidis et al. [29]), and the ‘TechRing’ by FirmTech [30]. However, these sensors all rely on ring-shaped strain gauge technology, which require pressure to be applied to the penile tissue to assess erection quality. In contrast, the approach presented in this study allows for the assessment of these parameters without the use of a pressure component, enhancing patient comfort. This innovative method has the potential to address the limitations of the RigiScan, which has raised concerns regarding the validity of measurements in overnight ambulatory settings [3].

While the current system demonstrates promise for ambulatory nocturnal erection detection and has proven component precision, further research is necessary. Initial proof-of-principle studies should focus on testing the key hypotheses outlined in this paper through feasibility studies in healthy men. These hypotheses include: (1) detecting detumescence using abdominal temperature measurements, (2) observing the absence of arterial pulse in the arteria cavernosa during full penile rigidity, and (3) detecting erections via accelerometry. The findings from these studies will provide critical insights for further system refinement and eventual clinical validation. Furthermore, the initial feasibility studies will contribute to a deeper understanding of the physiological mechanisms underlying ED.

The clinical applications of The Potential Sensor could extend beyond ED diagnosis to include monitoring recovery following prostatectomy in patients with prostate cancer. If sensor miniaturization can be achieved, the system could also be explored for use in research on female sexual dysfunction. Additionally, the use of readily available sensor components may enhance affordability, potentially enabling the system to become a point-of-care test accessible in pharmacies. This would represent a significant advancement toward promoting global acceptance of ED and reducing the stigma associated with the condition.

## Data Availability Statement

The data generated and analyzed during this study can be obtained from the corresponding author on reasonable request.

## Acknowledgements

Not applicable

## Author contributions

HJT conceived and designed the work, acquired data, and played an important role in interpreting the results. She drafted the manuscript, approved the final version and agreed to be accountable for all aspects regarding the accuracy and integrity of the work.

JTWB conceived and designed the work, acquired data, and played an important role in interpreting the results. She revised the manuscript, approved the final version and agreed to be accountable for all aspects regarding the accuracy and integrity of the work.

JGJK designed the work, acquired data, and played an important role in interpreting the results. He revised the manuscript, approved the final version and agreed to be accountable for all aspects regarding the accuracy and integrity of the work.

LIS played an important role in interpreting the results. She revised the manuscript, approved the final version and agreed to be accountable for all aspects regarding the accuracy and integrity of the work.

JJHB conceived and designed the work and played an important role in interpreting the results. He revised the manuscript, approved the final version and agreed to be accountable for all aspects regarding the accuracy and integrity of the work.

## Ethical Approval

The design and development of this proof-of-principle study did not require ethical approval. The Declaration of Helsinki was followed in the establishment and conduction of this study.

